# A novel interface for display of physiological data from human patient simulators

**DOI:** 10.1101/2025.09.02.25334904

**Authors:** R Roberts, R Russmann, L Cherrington, X Wang, A Cowen, D Morgan, R Helyer

## Abstract

Patient Simulators 1.0 is a novel display interface that provides an alternative means of showing real-time changes in physiological variables derived from human patient simulators. It is designed for teachers and learners of the sciences for whom showing variables on simulated clinical monitors may be less relevant. It allows import of comma separated data sets, and their display using customisable gauges and trendlines. Data from variables not normally shown on simulated patient monitors, e.g. pH, and data from variables that are not produced during the simulation but can be derived, e.g. stroke volume, can be displayed. It also allows presentation of data produced during simulation sessions carried out elsewhere without the need for proprietary simulation software. This interface may assist in wider adoption of the use of simulated data in teaching the basic science underpinning health and disease.

## Background

The application of human patient simulators to the teaching and learning of principles of physiology that underly health and disease is gaining interest (Harris et al, 2011; Waite et al, 2013; Helyer & Dickens, 2016; Beilby-Clarke, 2014; Bintley et al, 2019; Johnson et al, 2019). Human patient simulators such as CAE iStan (CAE Healthcare, USA) and Laerdal SimMan (Laerdal, Norway) are manikin-based and operate using physiological models of varying complexity. Although typical applications of simulation are to the learning of clinical skills and processes in healthcare, the underlying physiological models that are used to show vital signs such as pulse rate, and status of physiological variables such as pH and blood gases, are a rich data source. These data can be used by learners to explore, in depth, the key scientific principles underlying disease such as blood loss (Lloyd et al, 2006) or environmental challenges such as ascent to altitude (Helyer et al, 2009).

Simulation-based scenarios are typically run in real-time in the presence of learners, but also store physiological data and logs of events that can be run offline or used to explore key events in a snapshot of the simulation. These can used in debriefing, a typical follow-up to clinical simulations, but can also be used in data handling exercises to explore integrated body mechanisms. Such exercises form the basis of the application of simulation to the teaching of key scientific principles, forming the bridge between clinical diagnosis and practice, and the sciences.

Despite this potential, there are obstacles to the use of these powerful learning tools for science education (Helyer & Dickens, 2016). One unexpected fundamental issue we have experienced has been a dislike by science faculty for the way physiological data are presented to learners. Designed for clinical training, variables tend to be presented using a simulated clinical monitor in the style familiar to healthcare workers (Figure 1A).Clinical monitors do not show the entire range of variables that might be of interest in understanding underlying physiology, such as blood gases and pH. This can be circumvented by providing ancillary materials such as blood gas charts or displaying data to learners using the simulation instructor interface (Figure 1B) – we have utilised both methods. However, although of great value to learners in health sciences, these interfaces do not necessarily display data in a form that might be familiar or engaging to students of sciences. In one example in our curriculum, a proposed simulation designed to explore pressure changes during heart-failure for final year students of physiological science was not adopted because it was not possible to clearly visualise pressure changes at key moments during the disease progression using available tools. Further, some key variables, for example stroke volume and total peripheral resistance, are not logged directly and therefore are not displayed during the simulation in real-time.

**Figure 1.**
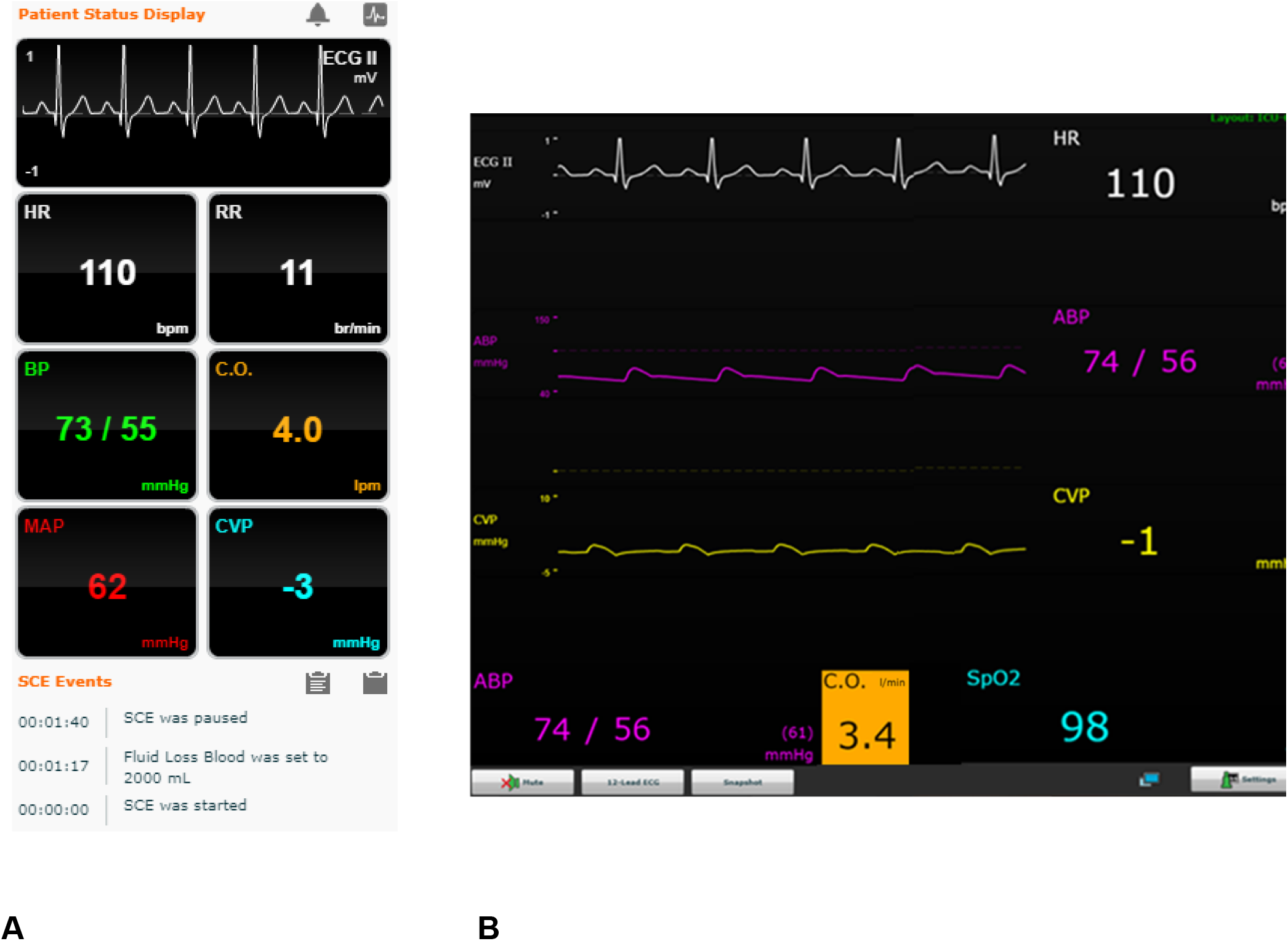
The interfaces available for display of data from physiological variables using the patient status display from the CAE Müse software interface (A) and CAE Touchpro simulated clinical monitor (B). All variables that are logged can be selected for display on the Müse interface, the simulated clinical monitor shows clinically relevant variables. Variables shown are simulated electrocardiogram (ECG) trace for lead II, heart rate (HR), respiratory rate (RR), blood pressure (BP/ABP), cardiac output (CO), mean arterial pressure (MAP), central venous pressure (CVP) and oxygen saturation of blood (SpO2).

We set out to design an alternative interface for the engaging, real-time display of simulated physiological variables for science students using data extracted from simulation data logs. This project was carried out by faculty that teach physiology and by second year undergraduate computer science students as a summative coursework project.

### Design of the application

Simulators that operate using software with physiological models such as CAE Müse for iStan log time-stamped data for selected variables during the simulation. They may also log events taking place, such as drug applications and other interventions. These data can be extracted from the simulation software either directly, or via series of intermediate steps, as delimited data tables that can be read by other software such as Microsoft Excel. Specifically, Müse allows export of data in the comma separated value (.csv) format.

The data visualisation application ‘Patient Simulators 1.0’ (PS1.0) was created using JavaFX (Oracle, USA) and HanSolo/Medusa gauges (available at Github.com). The application is a Java executable (jar) file.

The interface is shown in Figure 2. Data and event log files are loaded as.csv format. Parameters to display are selected via the ‘Headers’ tab. The display format is set by ‘gauge type’. Maximum and minimum values can be set for each chosen variable as well as coloured regions to indicate normal (green), borderline (amber) and abnormal (red) ranges for each one. The option of a cylinder style gauge was incorporated that allows values for variables to be displayed as a percentage of maximal values. The option of trend-line graphs were also incorporated. The application can be run in real-time or up to 8x and can be paused and replayed. Files may be saved as.sim files, and reloaded to retain the selected parameters and styles

**Figure 2.**
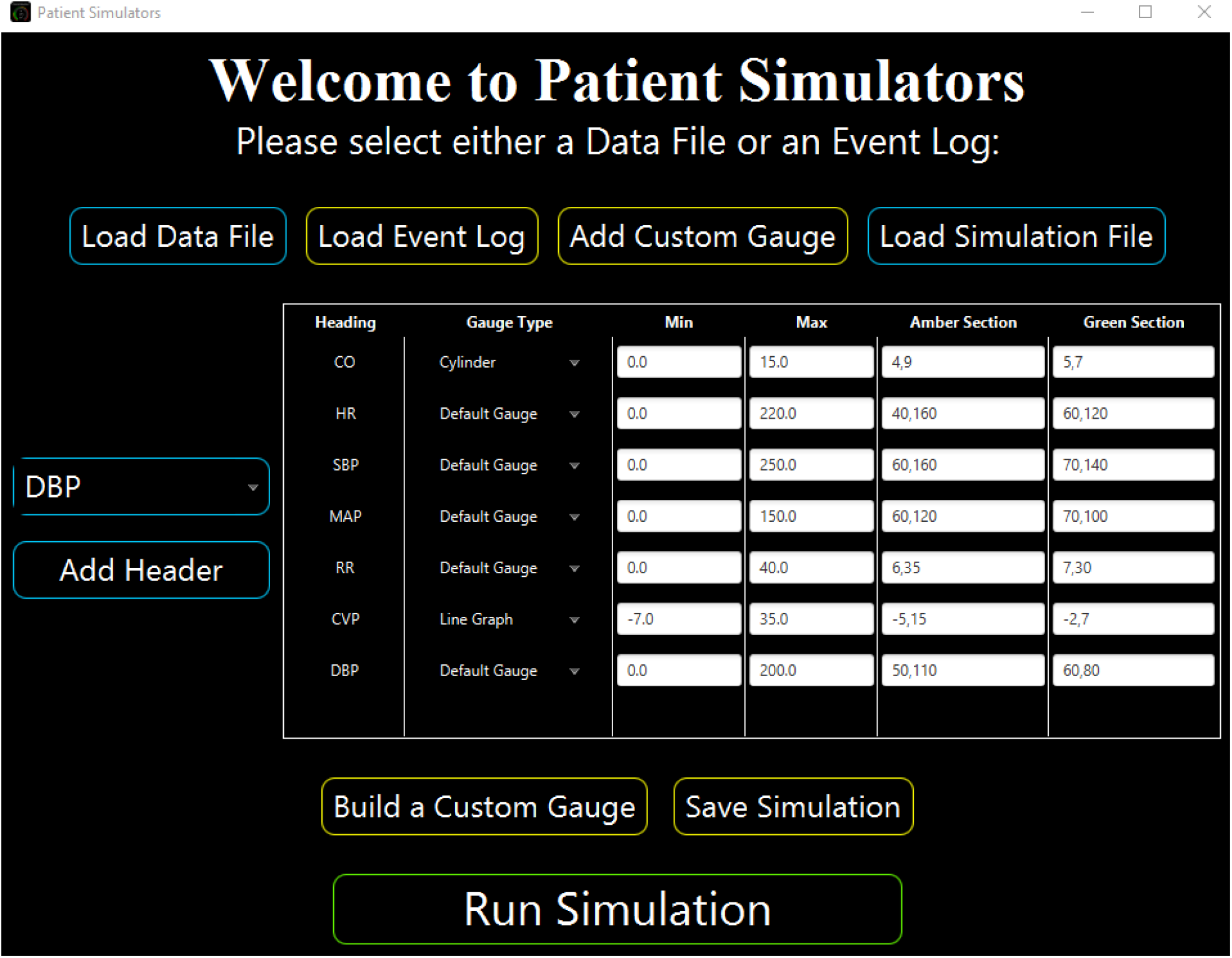
The Patient Simulators 1.0 interface. Data files and event logs in.csv file format are loaded, and physiological variables selected for display using ‘add header’. Gauge types can be selected from pre-defined options or customised. Minimum and maximum values for each gauge are set and ranges defined. Configurations can be saved as.sim files and reloaded

### An example of a simulation displayed using Patient Simulators 1.0

An example of the data display produced using PS1.0 is shown in Figures 3A and 3B. Data were derived from a simulation designed to demonstrate the physiological response to increasing levels of hypovolemia in a healthy adult male with the learning outcome to be able to explain the operation of the baroreceptor reflex. Students observe changes in values of key variables in real-time and note down values for subsequent analysis. In this example, data are shown in a variety of gauge and chart formats for illustrative purposes (Fig 3A) with approximations to normal, borderline and abnormal ranges. The cylinder gauge in this example represents the percentage of maximal CO at each time point. Line graphs may be shown in order to emphasise trends in real time for selected parameters. The display is completely customisable - any parameter can be displayed in any format. Figure 3B shows data for the same variables but after removal of two litres of whole blood volume (approximately 30% of the blood volume for a healthy adult male) later in the simulation. Gauges can be seen to be in different positions, their transit having occurred in real-time or at the selected running speed.

**Figure 3.**
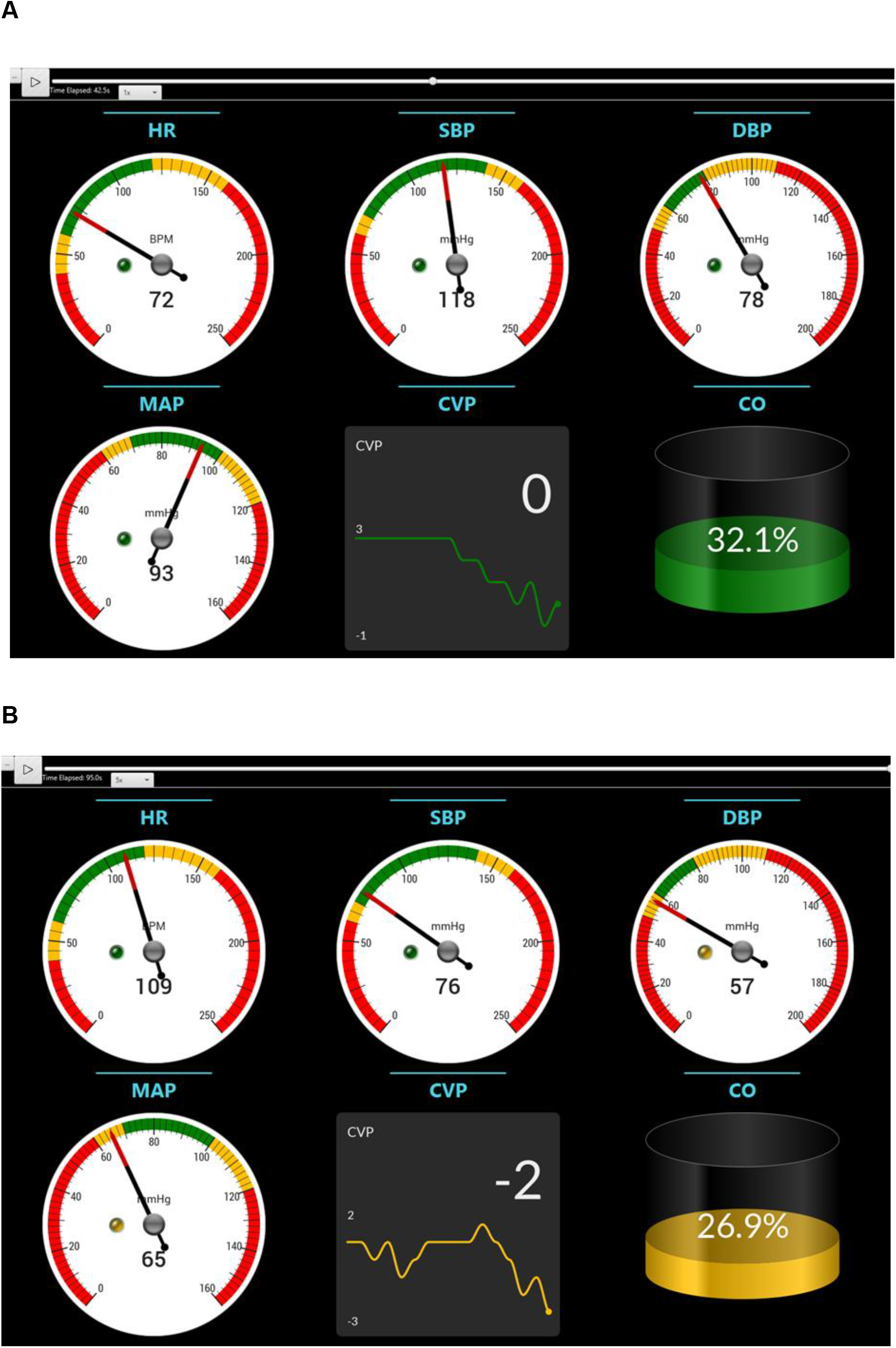
Snapshots of the Patient Simulators 1.0 display early and later in a simulation designed to explore the physiological mechanisms operating during blood loss. **A** shows values for a normal healthy individual early in the simulation and **B** after a simulated loss of 2 l of blood. In this configuration, gauges show heart rate (HR), systolic blood pressure (SBP), diastolic blood pressure (DBP) and mean arterial pressure (MAP). The line chart indicates the trend in central venous pressure (CVP) and the cylinder shows cardiac output as a percentage of maximal. Colours and coloured regions indicate normal (green), borderline (amber) and abnormal (red) ranges. Note values and ranges are for illustrative purposes only and are not intended to be clinically accurate.

### Potential applications and further developments

We set out to develop a display interface for physiological data derived from human patient simulators that shows changes in variables. PS1.0 allows customised display of data from variables in real-time and allows files to be saved for repeated use by teachers and learners offline without the need for proprietary simulation software.

It has several powerful features. First, it can show data in a gauge or trend format rather than viewing values as changing digits or repeating waveforms that do not show trends. Second, it can show data for any chosen variable from a simulation, including those typically not available on simulated clinical monitor interfaces, such as pH. Third, manipulation of the data file of simulated data (e.g. using Microsoft Excel) allows display of data for variables not produced directly by the simulation software. For example, data for stroke volume that are not produced using CAE Müse can be calculated and added into the data file for display. Indeed, any data from any source can be added for display if time-stamped and in an appropriate delimited file format. Finally, a feature that may aid adoption of the use of data high-fidelity simulation in science teaching is the ability to display data sets from simulations carried out elsewhere. For example, using PS1.0, data produced by a clinical simulation in a hospital setting elsewhere could be used in sciences teaching without the need for expensive proprietary simulation software – licences are often linked to individual simulator hardware.

PS 1.0 has advantages over other potential ways of displaying data. Trendlines can be shown in software such as Microsoft Excel, but as static graphs not showing changes in values in real-time. Programmes such as Matlab can produce displays of changing variables in real-time but require coding skills and the interface is more complex. PS1.0 is user friendly and requires no detailed knowledge of data manipulation.

Future developments planned include exploring the potential for displaying data live or with a small-time delay during a simulation rather than after a simulation. We are exploring the potential for use with data from sources such as clinical devices. It is intended Patient Simulators 1.0 will be made available open source to educational partners.

## Conclusion

PS1.0, designed for use by teachers and learners of science, is a novel interface allowing display of real-time changes in physiological variables generated using high-fidelity simulation.

## Data Availability

All data produced in the present work are contained in the manuscript

## Acknowledgements

The authors would like to thank the School of Computer Science University of Bristol for approving this collaborative undergraduate project. We thank Drs Lauren Goodhead and Phil Langton (University of Bristol) for discussions around requirements for visualisation of data for scientists. We thank Mr Peter Dickens, (Kliniskt Träningscentrum, Akademiska Sjukhuset, Uppsala, Sweden) for information on the workflow related to alternative simulators.

## Declarations

This project was solely funded by the University of Bristol. There are no conflicts of interest. There were no ethical considerations (simulated data).

